# Differential, not uniform, elevation of plasma neurofilament light and glial fibrillary acidic protein across acute psychiatric presentations: implications for biomarker interpretation in psychiatry

**DOI:** 10.1101/2025.11.19.25340633

**Authors:** Dhamidhu Eratne, Matthew JY Kang, Charles B Malpas, Samantha M Loi, Alexander F Santillo, Henrik Zetterberg, Naveen Thomas, Ian Everall, Chad Bousman, Christos Pantelis, Kruti Patel, Jeffrey Smith, Michael Christie, Cherie Chiang, Dennis Velakoulis, The MiND Study Group

**Author notes:** **Corresponding author:** Dhamidhu Eratne. Statistical analysis conducted by: Dr Dhamidhu Eratne.

## Abstract

**Objective:** To investigate plasma neurofilament light chain (NfL) and glial fibrillary acidic protein (GFAP) levels across a range of acute psychiatric disorder presentations, and to determine whether elevations are specific to certain diagnoses.

**Methods:** We analysed biobanked lithium heparin plasma samples from 121 patients with acute psychiatric presentations (including mania, psychosis, alcohol use disorder, anorexia nervosa, depression, and adjustment disorder), and 59 healthy controls. NfL and GFAP were measured using Simoa assays. Group differences were compared via bootstrapped general linear models adjusted for age, sex, and weight.

**Results:** Among 180 participants, adjusted NfL was significantly elevated compared with controls in alcohol use disorder (β=1.32, p=0.002), anorexia nervosa (β=0.86, p<0.001), mania (β=0.44, p=0.015), and psychosis (β=0.34, p=0.034), but not in depression or adjustment disorder. Unadjusted NfL was highest in alcohol use disorder (mean 43 pg/mL) and anorexia nervosa (20 pg/mL) versus controls (9 pg/mL). Adjusted GFAP was elevated only in alcohol use disorder (β=1.01, p=0.024).

**Conclusions:** Plasma NfL and GFAP are not uniformly elevated across psychiatric disorders, but show a disorder-specific pattern, with the largest elevations in alcohol use disorder and anorexia nervosa and more modest elevations in mania and psychosis. These findings support nuanced, biologically grounded, diagnosis- and stage-specific interpretation of these biomarkers in psychiatry, and caution against interpreting a single elevated value as evidence of neurodegeneration. Because some acute psychiatric presentations show elevations approaching the neurodegenerative range, control-derived reference ranges may be insufficient when distinguishing neurodegeneration from primary psychiatric disorders, supporting the need for diagnosis- and state-specific reference data.

## Introduction

Blood biomarkers that reflect central nervous system pathological processes, such as neurofilament light chain protein (NfL, a marker of neuronal injury) and glial fibrillary acidic protein (GFAP, a marker of glial activation and neuroinflammation), are increasingly recognised for their diagnostic and broader value in neurodegenerative disorders. More recently, there has been growing interest in whether these markers may be relevant in primary psychiatric disorders (PPD), particularly in severe or acute presentations where there may be neurobiological changes that overlap with neurodegeneration or neuroinflammation (Abu-Rumeileh et al., 2023; Bavato, Barro, et al., 2024; Bavato, Seifritz, et al., 2024; Dai et al., 2022). The validation and use of such blood tests could facilitate improved understanding of underlying biological processes, help identify unique subtypes, and serve as markers of treatment response and poorer prognosis. Given the increasing awareness of this association between diverse psychiatric disorders and dementia (Baudouin et al., 2025; Livingston et al., 2024), they may also enable early identification of individuals at greater risk for neurodegeneration and dementia.

Several studies have demonstrated elevated plasma NfL in bipolar affective disorder (BPAD) depression (Aggio et al., 2022; Kang, Eratne, et al., 2025), major depressive disorder (MDD) (Bavato et al., 2021), and psychotic disorders (Ceylan et al., 2023; Rodrigues-Amorim et al., 2020). However, findings have not been consistent (Squitti et al., 2024), with other studies not finding elevated levels in BPAD (Queissner et al., 2024), MDD (Eratne, M Kang, et al., 2024), or treatment-resistant schizophrenia and first-episode psychosis (Eratne et al., 2022; Eratne, M Kang, et al., 2024; Kang et al., 2024). A recent meta-analysis found elevated NfL levels in bipolar disorder, but not major depressive disorder (Kang, Grewal, et al., 2025). NfL and GFAP have been found to be elevated in bipolar depression (Kang, Eratne, et al., 2025) and anorexia nervosa (Doose et al., 2023; Hellerhoff et al., 2021; Nilsson et al., 2019). Neither biomarker was elevated in treatment-resistant schizophrenia (Eratne, MJY Kang, Lewis, Dang, Malpas, Ooi, et al., 2024).

A major limitation in the literature has been the lack of research in different phases of PPD, particularly acute phases of illness. Recent studies of NfL in a large group of psychiatric emergency presentations found elevated levels across a diverse range of broad diagnostic categories, from personality to anxiety to mood disorders, however, their mood disorders cohort was not further characterised (i.e. in to unipolar vs bipolar disorder), and did not provide any findings in patients with mania (Light et al., 2024; Zarglayoun et al., 2025). Ceylan et al. investigated adolescent patients with bipolar mania, however analyses were not adjusted for important covariates such as weight, age, and sex, known to affect blood biomarker levels. To our knowledge, only two studies have reported biomarkers in mania, n=4 in each study. (Al Shweiki et al., 2019; Steinacker et al., 2021), but only Steinacker et al. reported findings in mania specifically (n=4; NfL and GFAP levels were not elevated).

Despite this growing literature, there remain several key unanswered questions. First, few studies have examined individuals during acute psychiatric presentations, when symptoms are most severe and biological changes such as neuronal injury and neuroinflammation may be most pronounced. Second, very few studies have focused on mania and acute psychosis, owing to recruitment challenges during the acute phases of illness. Third, potential confounders, such as age and body weight, have not always been adequately addressed. Finally, no studies to our knowledge have yet described GFAP levels in a diverse range of acute psychiatric disorder presentations.

This study aimed to examine plasma NfL and GFAP levels in individuals presenting to hospital with a range of acute psychiatric disorders, including mania, psychosis, anorexia nervosa, alcohol use disorder, depression, and adjustment disorder, and compare these to levels in healthy controls. We hypothesised that NfL and/or GFAP would be elevated in some acute psychiatric presentations — particularly mania and psychosis – but not across all diagnostic groups – potentially reflecting differing degrees of neuronal injury and neuroinflammation in acute phases of severe disorders compared to acute presentations of less severe disorders.

## Methods

### Recruitment and sample analysis

Patients in this study had been admitted with acute psychiatric presentations to acute settings at The Royal Melbourne Hospital (RMH), including the emergency department, an acute adult psychiatry inpatient unit, and an eating disorder inpatient unit. The broad admission category was reviewed, and patients with manic, depressed, psychotic, anorexia nervosa and other psychiatric presentations were identified. People admitted with alcohol use disorder were identified as a comparator, positive control group, given the evidence of NfL and GFAP elevations in this alcohol use disorder (Hou et al., 2025; Huang et al., 2024; Requena-Ocaña et al., 2023). Routine admission pathology samples from the identified inpatients were retrieved, and de-identified prior to analysis. Normal renal function, defined as an estimated glomerular filtration rate (eGFR) within the normal reference range on routine admission pathology, was an eligibility criterion; samples from patients with renal impairment were excluded, given the known influence of reduced renal clearance on plasma NfL and GFAP concentrations. Waiver of consent was approved by the hospital ethics committee for the use of routine de-identified blood samples and data for the development of in-house biomarker testing (HREC/65332/mh-2020). Aliquots were stored at −80C, and analysed for NfL and GFAP using N2PB kits on Quanterix Simoa HD-X analysers. The primary discharge diagnosis was extracted from file review and was categorised into one of the following: adjustment disorder, alcohol use disorder, anorexia nervosa, depression, mania (bipolar mania), other PPD, psychosis (schizophrenia spectrum disorders). Sex, weight and age were linked to each sample episode.

The control group consisted of unrelated, age, sex and sociodemographic matched healthy controls recruited from the general community, through the CRC psychosis study, as described in detail previously (Bousman et al., 2019; Eratne et al., 2022; Mostaid et al., 2017).

### Statistical analyses

All statistical analyses were performed in R version 4.5.0 (2025-04-11). A number of cases had missing values for weight (n=46). Due to weight having a significant influence on plasma biomarkers, missing weight data was imputed using multiple imputation by chained equations (MICE) with predictive mean matching, based on age, sex, and diagnostic group (mice package), with imputed values replacing missing ones in the main dataset. General linear models (GLMs) were fitted separately for logliltransformed plasma NfL and GFAP concentrations, with diagnostic group as the primary predictor and age, sex, and weight as covariates. Model coefficients and confidence intervals were estimated using nonlilparametric bootstrapping with 10,000 replicates (parameters package), due to small sample sizes and to ensure robust inference. Estimated marginal means (EMMs) were obtained using the emmeans package to calculate adjusted mean log values for each diagnostic group. Biomarker data from the control group were analysed in a separate batch using plasma EDTA samples using NfL and GFAP single-plex kits and Simoa SR-X analysers. Using data from samples that were analysed in both batches (n=72 from a separate study), we were able to determine a strong linear correlation between batches (Pearson’s correlations 0.97 and 0.96 for NfL and GFAP, respectively) and harmonise the control data.

## Results

The final study cohort included 180 participants with the following primary diagnoses: adjustment disorder (n=9), alcohol use disorder (n=10), anorexia nervosa (n=21), mania (n=17), depression (n=15), other PPD (n=13), psychosis (n=36), and controls (n=59). As illustrated in Table 1, there were age, weight, and sex differences between the groups. People with anorexia nervosa were the youngest (mean age 25 years). People with alcohol use disorder and depression were older (mean 48 years and 44 years, respectively), compared to the other groups (means 32-40 years). Imputed weights were similar to weights without imputation. Anorexia nervosa had the lowest weight (mean 43kg). Adjustment disorder and mania had higher weights compared to controls (87kg and 82kg, respective, vs 76kg).

**Table 1.** Study cohort details and biomarker levels. Data is Mean (SD) or n / N (%) z: Adjusted levels are estimated marginal means derived from general linear model with age, sex, and imputed weight as covariates (see Supplementary Material for further details) GFAP: glial fibrillary acidic protein; NfL: neurofilament light chain protein

| Characteristic | N | Adjustment disorder N = 9 | Alcohol use disorder N = 10 | Anorexia N = 21 | Depression N = 15 | Mania N = 17 | Other PPD N = 13 | Psychosis N = 36 | Control N = 59 |
| --- | --- | --- | --- | --- | --- | --- | --- | --- | --- |
| Age (years) | 180 | 40 (7) | 48 (21) | 25 (8) | 44 (19) | 32 (10) | 36 (19) | 39 (14) | 40 (11) |
| Sex | 180 |  |  |  |  |  |  |  |  |
| F |  | 4 / 9 (44%) | 4 / 10 (40%) | 20 / 21 (95%) | 7 / 15 (47%) | 9 / 17 (53%) | 9 / 13 (69%) | 21 / 36 (58%) | 22 / 59 (37%) |
| M |  | 5 / 9 (56%) | 6 / 10 (60%) | 1 / 21 (4.8%) | 8 / 15 (53%) | 8 / 17 (47%) | 4 / 13 (31%) | 15 / 36 (42%) | 37 / 59 (63%) |
| Weight (kg) | 134 | 85 (27) | 75 (19) | 44 (9) | 68 (13) | 83 (23) | 70 (19) | 75 (27) | 75 (11) |
| Missing |  | 6 | 2 | 1 | 7 | 4 | 6 | 15 | 5 |
| Weight (imputed) | 180 | 87 (20) | 78 (27) | 43 (9) | 67 (13) | 82 (22) | 68 (16) | 77 (23) | 76 (12) |
| NfL (pg/mL) | 180 | 7 (2) | 43 (49) | 20 (32) | 9 (6) | 10 (7) | 13 (11) | 13 (10) | 9 (7) |
| GFAP (pg/mL) | 180 | 86 (41) | 180 (127) | 127 (79) | 107 (67) | 92 (60) | 101 (61) | 99 (113) | 73 (34) |
| Log10NfL | 180 | 0.84 (0.14) | 1.40 (0.47) | 1.12 (0.32) | 0.89 (0.28) | 0.94 (0.23) | 0.98 (0.40) | 1.01 (0.31) | 0.93 (0.17) |
| Log10GFAP | 180 | 1.90 (0.19) | 2.13 (0.39) | 2.03 (0.27) | 1.96 (0.24) | 1.88 (0.29) | 1.93 (0.27) | 1.87 (0.30) | 1.82 (0.20) |
| Adjusted logNfL<sup>z</sup> |  | 0.88 | 1.31 | 1.17 | 0.79 | 1.04 | 1.02 | 1.01 | 0.91 |
| Adjusted logGFAP<sup>z</sup> |  | 1.94 | 2.09 | 1.99 | 1.91 | 1.95 | 1.94 | 1.87 | 1.82 |

### NfL in acute psychiatric presentations

As demonstrated in Table 1 and Figure 1, raw, unadjusted NfL levels were higher in alcohol use disorder (mean 43pg/mL) and anorexia nervosa (mean 20pg/mL), compared to controls (mean 9pg/mL). Descriptively, unadjusted levels in other groups were slightly higher compared to controls (psychosis 13pg/mL, other PPD 13pg/mL, mania 10pg/mL), or similar to controls (depression, adjustment disorder).

**Figure 1.**
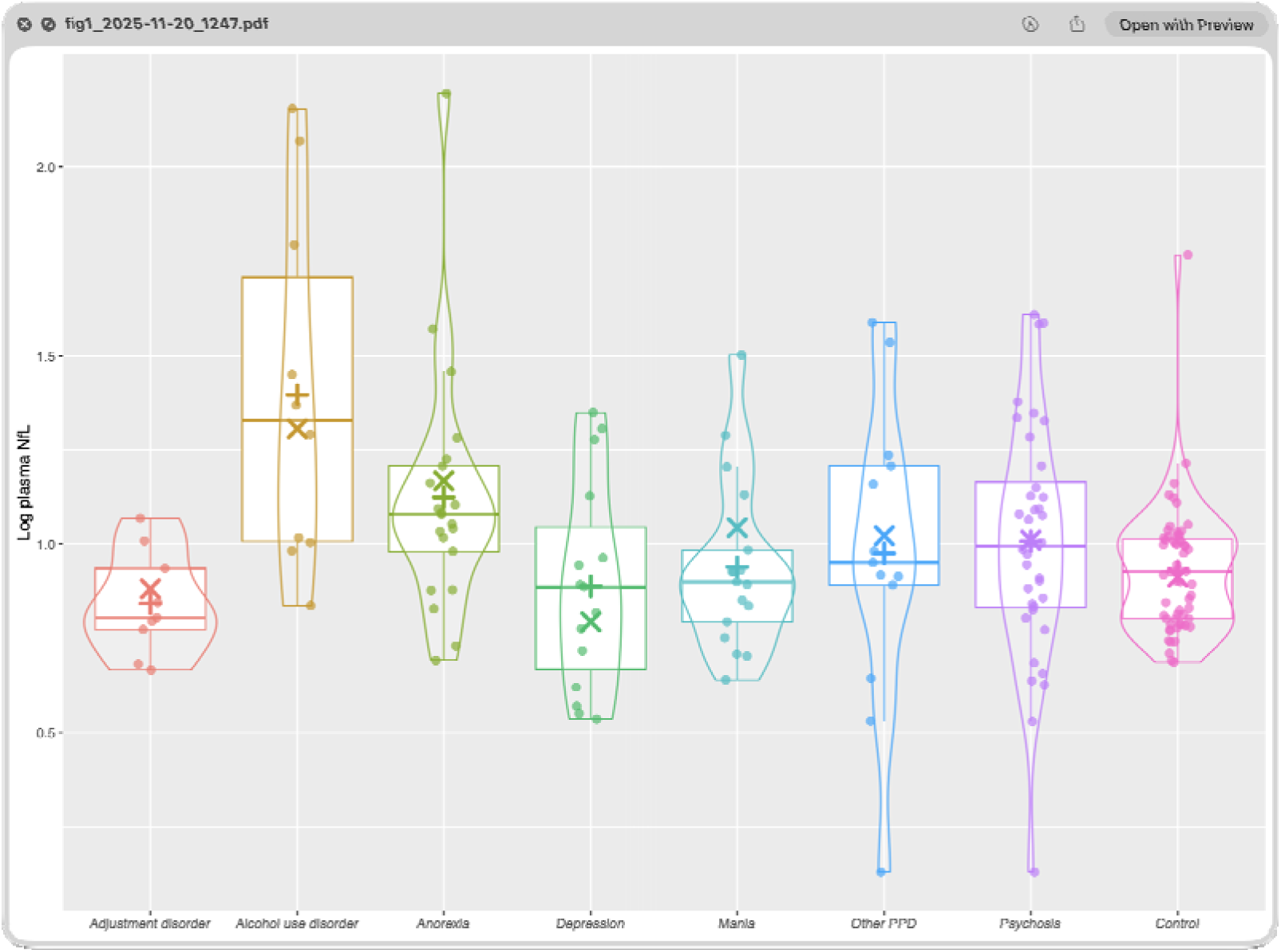
Plasma neurofilament light chain protein (NfL) levels in acute psychiatric disorder presentations and controls. + : unadjusted mean level X = adjusted mean level

To account for differences between groups, standardised bootstrapped GLMs with age, sex, and weight as covariates were performed, and adjusted log means derived for additional description (Table 1, Figure 1, and further details in Supplementary Material). These demonstrated markedly elevated NfL levels compared to controls in alcohol use disorder (adjusted mean log NfL 1.31 vs 0.91; β=1.32 95%CI:[0.53, 2.20], p=0.002) and anorexia nervosa (1.17 vs 0.91; β=0.86 [0.43, 1.32], p<0.001). NfL levels were also elevated in mania (1.04 vs 0.91; β=0.44 [0.08, 0.83], p=0.015), and psychosis (1.01 vs 0.91; β=0.34 [0.03, 0.66], p=0.034). None of the other groups were statistically different compared to controls.

Comparing only PPD to each other, with depression as the reference group, NfL levels were elevated in alcohol use disorder (1.31 vs 0.79; β=1.53 [0.80, 2.30], p<0.001) and anorexia nervosa (1.17 vs 0.79; β=1.15 [0.61, 1.64], p<0.001), mania (1.04 vs 0.79; β=0.81 [0.33, 1.27], p=0.002), psychosis (1.01 vs 0.79; β=0.68 [0.28, 1.07], p=0.003), and other PPD (1.02 vs 0.79; β=0.73 [0.12, 1.33], p=0.022). NfL levels between adjustment disorder and Depression were not statistically different.

### GFAP in acute psychiatric presentations

Unadjusted GFAP levels were markedly elevated in alcohol use disorder (180pg/mL) and anorexia nervosa (127pg/mL), compared to controls (73pg/mL), with less pronounced elevations in other groups (92-107pg/mL, Table 1 and Figure 2).

**Figure 2.**
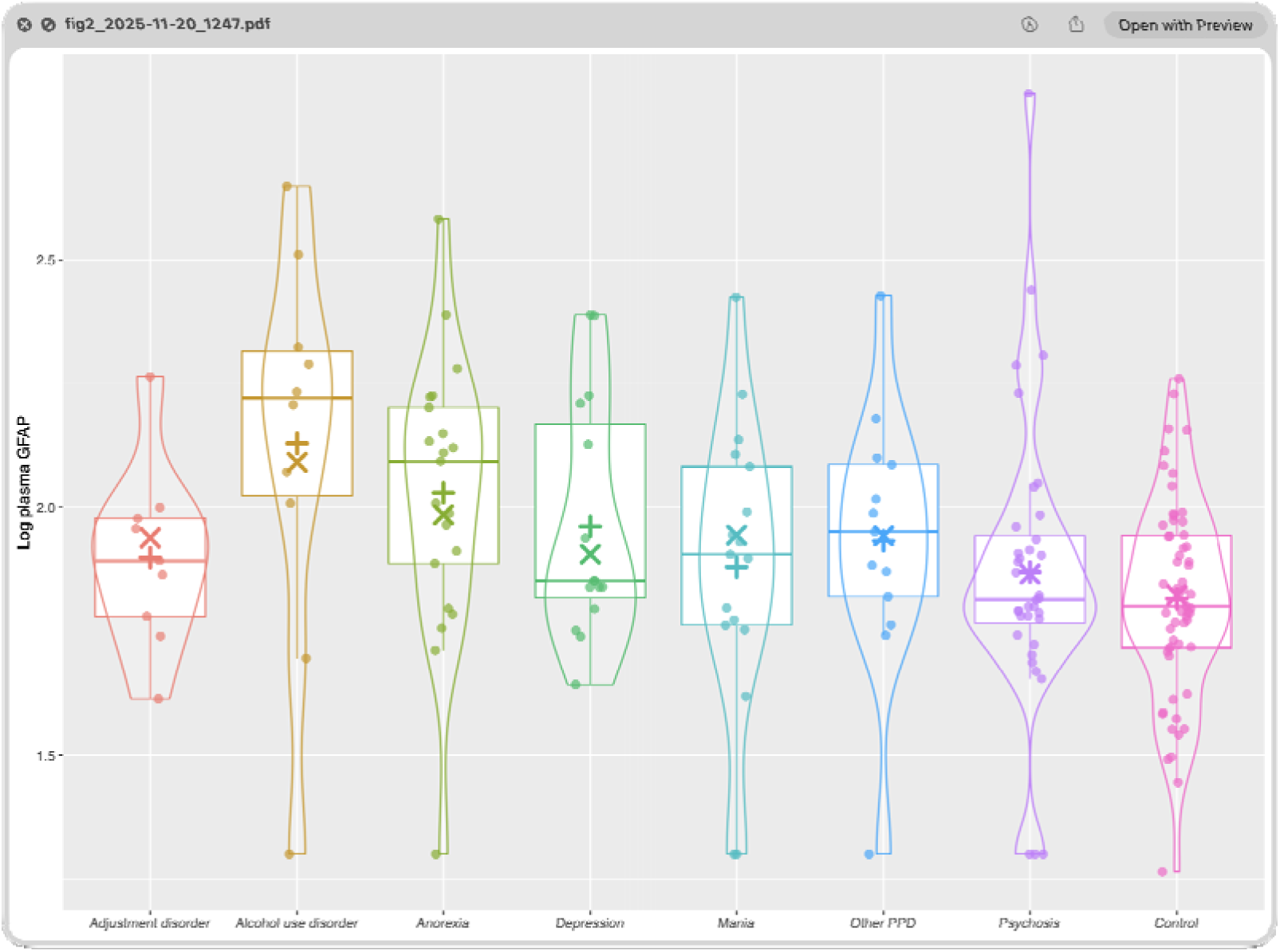
Plasma glial fibrillary acidic protein (GFAP) levels in acute psychiatric disorder presentations and controls. + : unadjusted mean level X = adjusted mean level

Adjusted log mean GFAP levels were only elevated in Alcohol use disorder compared to controls (2.09 vs 1.82; β=1.01 [0.16, 1.80], p=0.024).

Comparing only PPD to each other, with Depression as the reference group, GFAP levels were not elevated in any groups compared.

## Discussion

This study investigated plasma NfL and GFAP levels in people with acute psychiatric presentations and alcohol use disorder. The main finding after adjusting for important covariates (age, sex, and weight), was of elevated NfL levels in several acute psychiatric disorder presentations: markedly elevated significant levels in alcohol use disorder and anorexia nervosa, with more modest but significant elevations in mania and psychosis. To our knowledge, no previous study has described NfL and GFAP levels in bipolar mania and acute psychosis in adults. These elevations may reflect neuronal or neuroaxonal changes during the acute phases of these severe psychiatric disorders, although the cross-sectional design precludes any conclusion about injury, mechanism, or causation. Furthermore, this study is consistent with prior reports of elevated NfL in alcohol use disorder and anorexia nervosa. Our study is the first to describe GFAP levels in a diverse range of acute psychiatric presentations, not finding elevated levels in any of the disorders (except alcohol use disorder) compared to controls, or compared to each other. Strengths of our study include the inclusion of patients who are traditionally extremely challenging to recruit, robust statistical and imputation methods to mitigate against smaller sample sizes and missing weight data, and adjust for important covariates. A further strength is that normal renal function was an eligibility criterion, which reduces the likelihood that the observed elevations were attributable to impaired renal clearance, a recognised confounder of plasma NfL and GFAP.

Our findings differ from recent large-scale studies such as Light et al. and Zarglayoun et al. (Light et al., 2024; Zarglayoun et al., 2025), who reported broad elevations in NfL and GFAP across nearly all major psychiatric categories, also including anxiety disorders and personality disorders. By contrast, our study did not find uniform elevations. NfL levels in adjustment disorder and depression were comparable to controls, and GFAP elevations were limited to alcohol use disorder. Our findings that peripheral markers of neuronal (NfL) and glial (GFAP) origin are elevated only in certain severe subgroups or clinical states, is more plausible in that it may more accurately reflect underlying biological processes. NfL elevations in mania and psychosis may reflect acute neurotoxicity associated with extreme mood dysregulation, sleep-wake disruption, and agitation. This interpretation is supported by longitudinal neuroimaging evidence showing that a greater number of manic episodes is associated with progressive cortical thinning, particularly in prefrontal regions (Abé et al., 2022).

GFAP elevations in alcohol use disorder are consistent with prior evidence of astroglial activation and blood-brain barrier permeability changes in this population (Nadler et al., 2025). Normal NfL and GFAP levels may reflect either the lower biological burden in states such as adjustment disorder, or in the case of depression the possibility that such changes emerge only in chronic, severe, recurrent illness. Importantly, the presence of normal biomarker levels in some psychiatric groups adds credence to the signal observed in others – elevated levels in severe psychiatric disorders like bipolar disorder, and normal levels in milder disorders like adjustment disorder – are biologically plausible.

Notably, the largest elevations occurred in alcohol use disorder and anorexia nervosa, conditions with well-established central nervous system effects, indicating that the assays detected biologically expected signal; the absence of elevation in depression and adjustment disorder is therefore unlikely to reflect insufficient assay sensitivity or power alone. If all groups had shown elevated NfL or GFAP, regardless of diagnosis or severity, it would raise concerns about the specificity or utility of these markers. Instead, our results suggest that blood-based biomarkers could potentially identify differences between psychiatric presentations, reflecting plausible underlying biological processes, and may even help identify neuronal- and inflammation-related subtypes within our current heterogeneous diagnostic categories.

These findings also have a specific implication for how NfL and GFAP are interpreted in practice. The clinically important question is rarely whether a patient differs from a healthy control, but whether their presentation reflects a neurodegenerative or a primary psychiatric disorder. Our data show that this distinction is complicated by differential, and sometimes substantial, elevations within PPD itself, most notably in acute alcohol use disorder and anorexia nervosa, where levels approached those seen in neurodegenerative disorders. Reference ranges derived from healthy controls, and even from chronic or stable PPD, may therefore be insufficient, or may misclassify, when applied to acute psychiatric presentations without regard to diagnosis and illness state. This extends our previous work using large reference cohorts and age-adjusted models to distinguish neurodegenerative from primary psychiatric disorders (Eratne et al., 2024), and is consistent with evidence that diagnostic cut-offs are age-dependent (Light et al., 2024). Establishing diagnosis- and state-specific reference data, across the psychiatric spectrum and across illness phases, is an important direction for future work and a prerequisite for translating these biomarkers into psychiatric and neuropsychiatric triage.

We have previously described elevated NfL levels in bipolar depression (Eratne, M Kang, et al., 2024; Kang, Eratne, et al., 2025). Together with this study’s finding of elevated levels in mania, and our previous finding of mildly elevated levels in bipolar depression, these results are consistent with NfL elevations across different phases of bipolar illness, although a within-subject longitudinal design would be required to confirm state-dependence. The finding of normal biomarker levels in depression in this study is similar to our previous findings of normal NfL levels in unipolar depression (Eratne, M Kang, et al., 2024). We have now also described NfL levels in all stages of psychosis ultra-high risk and first-episode (Kang et al., 2024), treatment-resistant (Eratne et al., 2022), and now acute psychosis. In another study, we found lower levels of NfL in younger people with TRS (Wannan et al., 2024). In contrast, in similarly aged people with acute psychosis in this study, we found elevated levels. This study’s findings of elevated levels in acute psychosis contrasts to our negative findings in other phases/stages of the illness, and might suggest dynamic changes and processes, with elevated NfL only during acute phases. This could be consistent with findings of cortical thinning in psychosis occurs in the first few years following onset (Cropley et al., 2017; Sun et al., 2009), with evidence from neuropathology of highest brain iron elevations during early stages of schizophrenia patients, consistent with higher oxidative stress load (Lotan et al., 2023). Future studies should in larger groups try and identify clusters that are driving these changes in the possibility that these different clusters have different underlying biological mechanisms.

It is important to note that the magnitude of elevation varied considerably by group. In mania and psychosis the elevations were modest (unadjusted NfL approximately 1.1- to 1.4-fold higher than controls), whereas alcohol use disorder and anorexia nervosa showed substantially larger elevations (approximately 2- to 5-fold on unadjusted levels). With the exception of alcohol use disorder, these elevations were of lower magnitude than the approximately 2.5-fold (around 250%) elevations typically seen in neurodegenerative disorders such as behavioural variant frontotemporal dementia (Eratne, MJY Kang, Lewis, Dang, Malpas, Keem, et al., 2024). While this is consistent with lower-magnitude elevations than those in neurodegenerative disorders, it is conceivable that subtle but recurrent or sustained elevations, over a longer period of time and/or with repeated episodes, could contribute to the recognised increased risk, among people with psychiatric disorders, of dementia later in life; this hypothesis would require longitudinal data to test (Baudouin et al., 2025; Livingston et al., 2024). Therefore, biomarkers such as NfL could play a role as an objective biomarker identify individuals at high risk of developing dementia, to lead to earlier interventions to reduce risk and delay or prevent dementia, and as a marker of treatment response and prognosis.

There are several important limitations to this study. This was a cross-sectional study, limiting causal inference and precluding any conclusions about longitudinal change or biomarker dynamics over time. In particular, although the acute timing of sampling is consistent with state- or phase-dependent elevations, the absence of within-subject longitudinal data means that state-dependence cannot be established directly and remains an inference from cross-sectional and cross-study comparisons. The waiver of consent ethics meant that while we could include patients who traditionally are extremely hard to recruit (e.g., acute manic and psychotic presentations), we had very limited data available to investigate for other important covariates and associations, such as disease duration, substance and medication use. While the manic and psychosis groups were comparable to or larger than other studies, sample sizes in each group were still relatively small, and it is possible that we were underpowered to detect true differences, reflected by several analyses trending towards significance. Unfortunately, we were unable to harmonise our data to be able to directly compare acute mania and psychosis, to other stages of bipolar and schizophrenia disorders from our previous studies. We used a separate control group and robustly harmonised data, due to not having a local control group with weight data. Our findings should therefore be interpreted with appropriate caution and warrant replication, and should prompt larger studies with important clinical (e.g., substance use, number of episodes, medication use), neuroimaging (e.g., atrophy, white matter disease) covariates. Longitudinal follow up and serial biomarkers will help answer the question of whether prognosis, treatment-response, and clinical response can be tracked and guided by NfL and GFAP.

To our knowledge, this is the first study to report elevated plasma NfL in a group of people with acute mania and psychosis, and to highlight differential NfL and GFAP changes across psychiatric disorders using carefully adjusted models. These findings challenge the idea that neurodegeneration-associated biomarkers are elevated in all psychiatric disorders, and instead suggest more nuanced, as well as stage specific roles. If replicated, these results could inform future biomarker-based tools to differentiate between psychiatric diagnoses, stratify severity, track treatment response, and identify individuals at risk of poor prognosis or neurodegenerative progression. Future work should explore longitudinal trajectories, incorporate neuroimaging or cognitive correlates, and include larger, phenotypically richer cohorts. Understanding whether these biomarker elevations persist, resolve, or predict outcomes will be critical to determining their utility in clinical psychiatry.

## Data Availability

All data produced in the present study are available upon reasonable request to the authors

## Acknowledgements and funding sources

The authors would like to thank all the participants, patients and their families, for their participation.

The corresponding author had full access to all the data in the study and had final responsibility for the decision to submit for publication.

This research was supported by Colonial Foundation. Funding was only for sample collection and biomarker analyses and did not influence results or interpretation. Dhamidhu Eratne is supported by NHMRC Ideas Grant 1185180.

Matthew Kang is supported by the Research Training Program Scholarship (stipend) from the Department of Psychiatry, University of Melbourne with contributions from the Australian Commonwealth Government and the Ramsay Health Research Foundation Translation Challenge.

HZ is a Wallenberg Scholar and a Distinguished Professor at the Swedish Research Council supported by grants from the Swedish Research Council (#2023-00356, #2022-01018 and #2019-02397), the European Union’s Horizon Europe research and innovation programme under grant agreement No 101053962, Swedish State Support for Clinical Research (#ALFGBG-71320), the Alzheimer Drug Discovery Foundation (ADDF), USA (#201809-2016862), the AD Strategic Fund and the Alzheimer’s Association (#ADSF-21-831376-C, #ADSF-21-831381-C, #ADSF-21-831377-C, and #ADSF-24-1284328-C), the European Partnership on Metrology, co-financed from the European Union’s Horizon Europe Research and Innovation Programme and by the Participating States (NEuroBioStand, #22HLT07), the Bluefield Project, Cure Alzheimer’s Fund, the Olav Thon Foundation, the Erling-Persson Family Foundation, Familjen Rönströms Stiftelse, Familjen Beiglers Stiftelse, Stiftelsen för Gamla Tjänarinnor, Hjärnfonden, Sweden (#FO2022-0270), the European Union’s Horizon 2020 research and innovation programme under the Marie Skłodowska-Curie grant agreement No 860197 (MIRIADE), the European Union Joint Programme – Neurodegenerative Disease Research (JPND2021-00694), the National Institute for Health and Care Research University College London Hospitals Biomedical Research Centre, the UK Dementia Research Institute at UCL (UKDRI-1003), and an anonymous donor. CP was supported by a National Health and Medical Research Council (NHMRC) L3 Investigator Grant (1196508) and NHMRC Program Grant (ID: 1150083). CRC Psychosis study was funded by a Department of Industry Co-operative Research Centre (CRC) grant (ID: 20090064; investigators included IE, CP).

## Appendix: collaborators/contributors

On behalf of others in The MiND Study Group:

## Supplementary material

### PPD vs controls with imputed weight – adjusted log means derived from GLMs

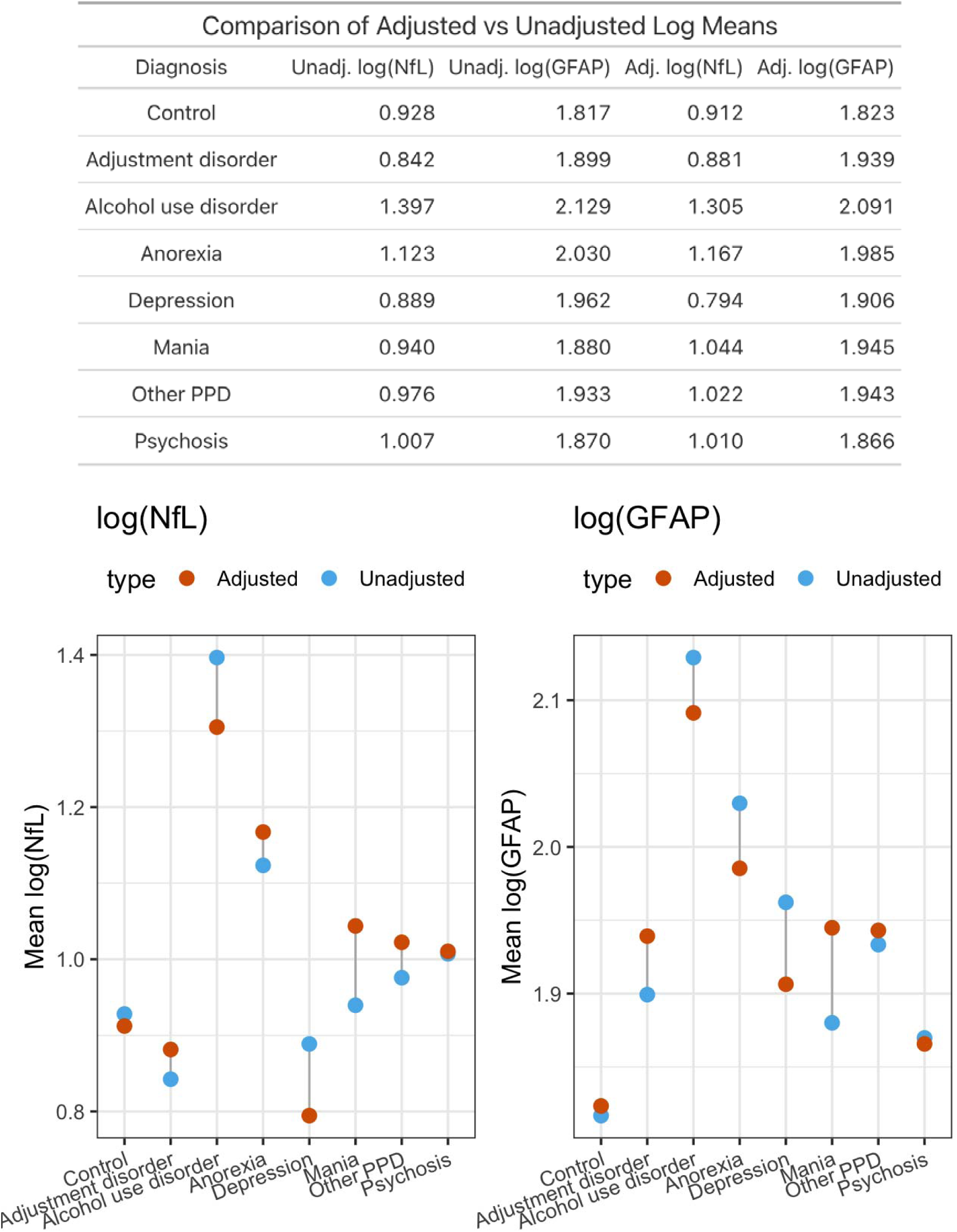

### PPD vs controls with imputed weight (reference = Controls) – GLM results

#### NfL

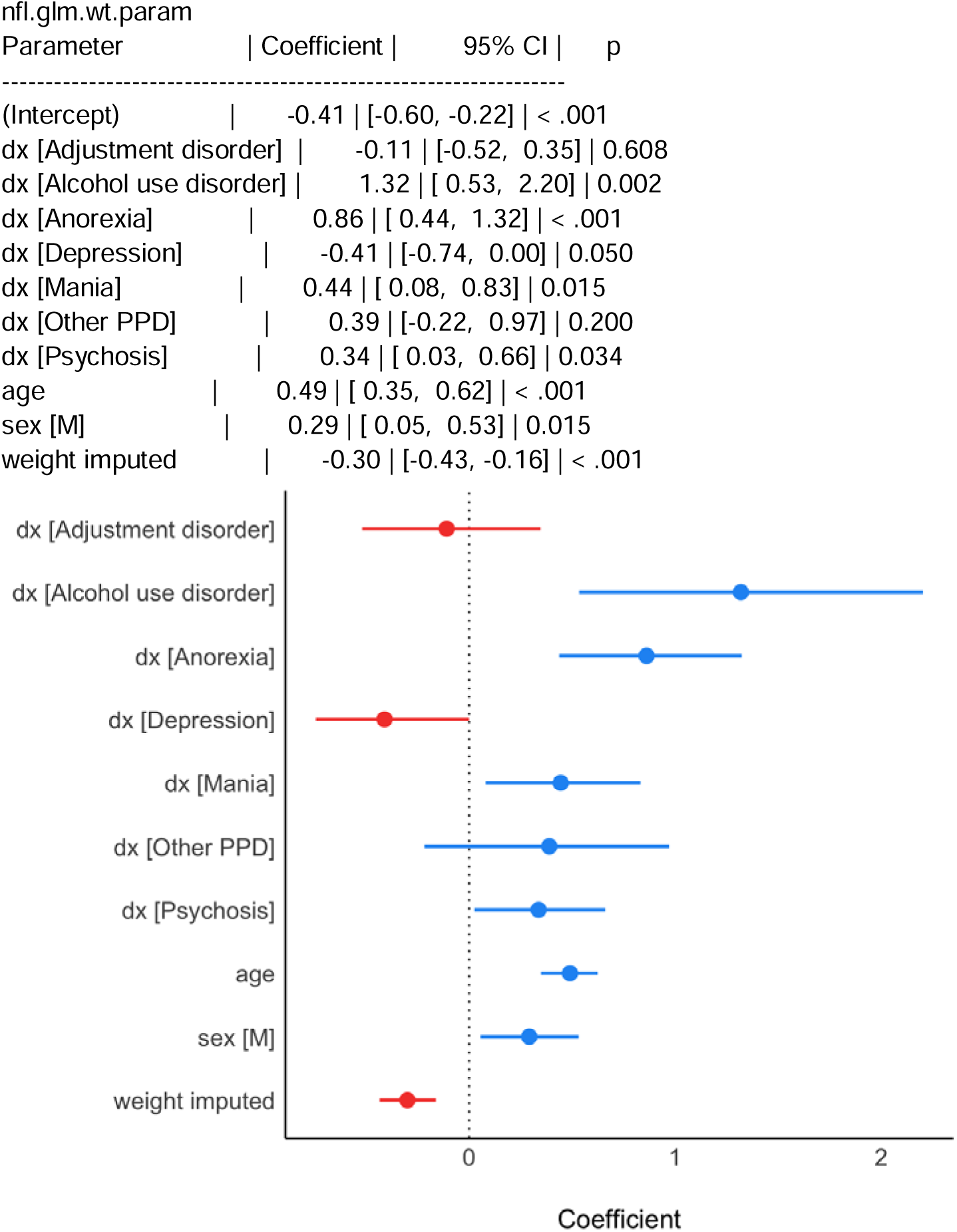

#### GFAP

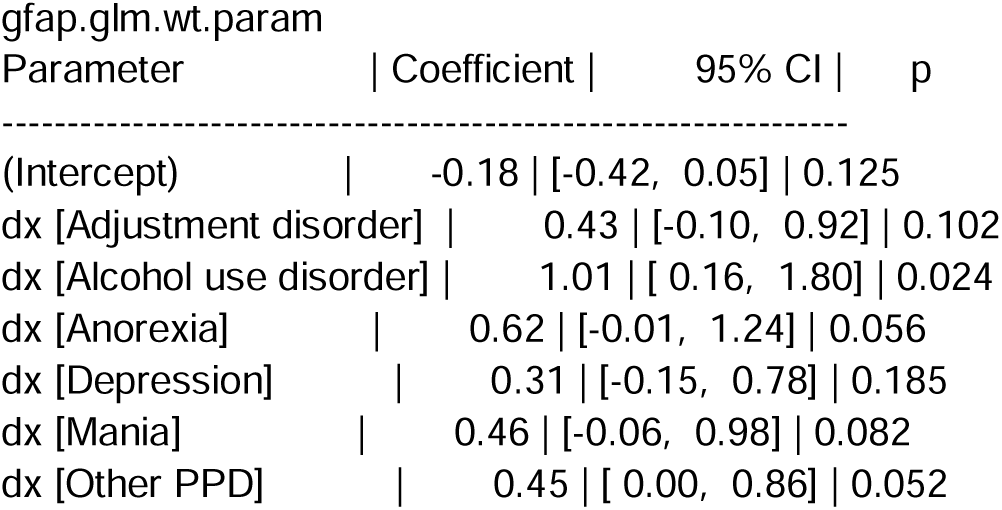
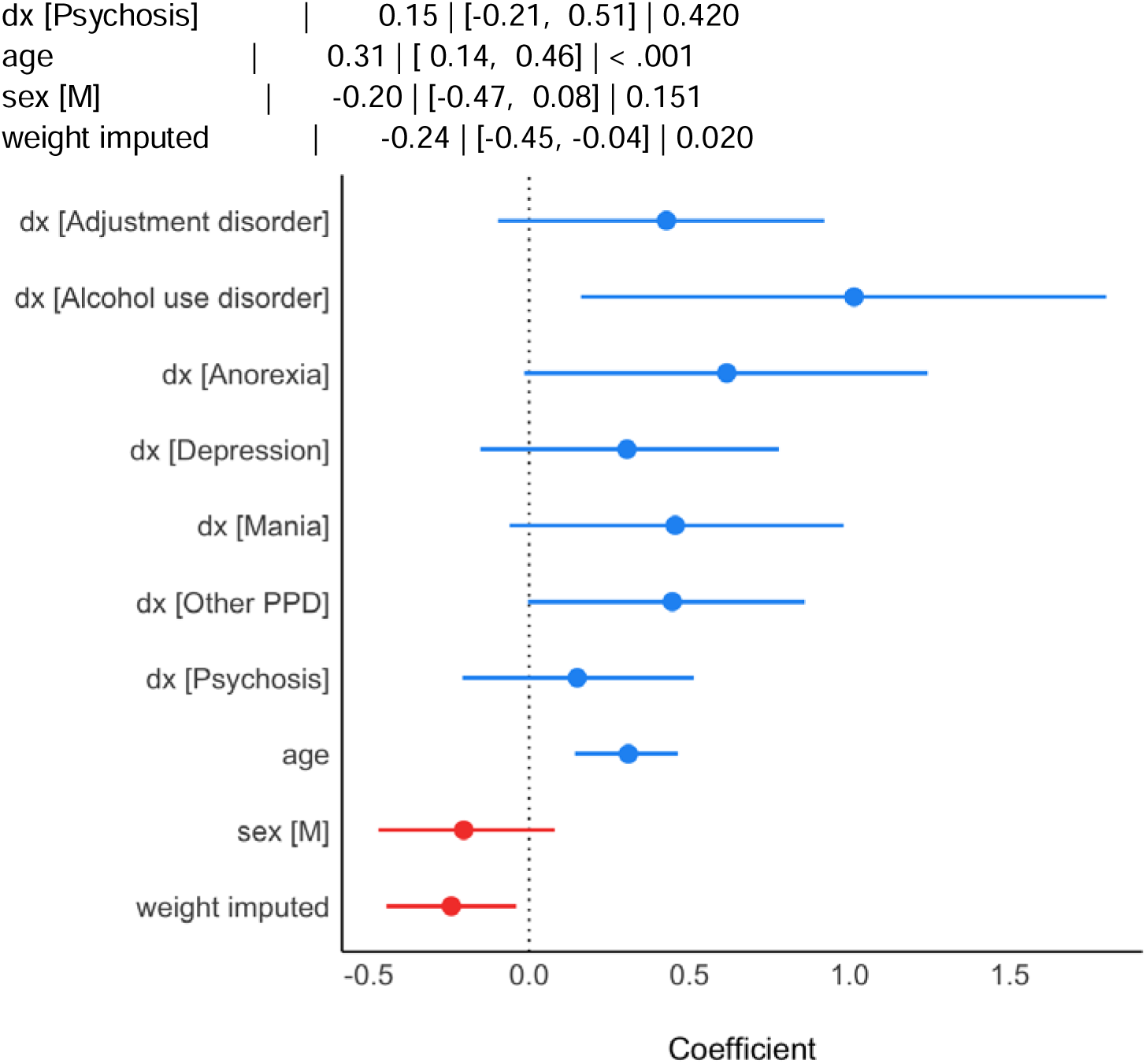

### PPDs compared to each other with imputed weights (reference = Depression) – GLM results

#### NfL

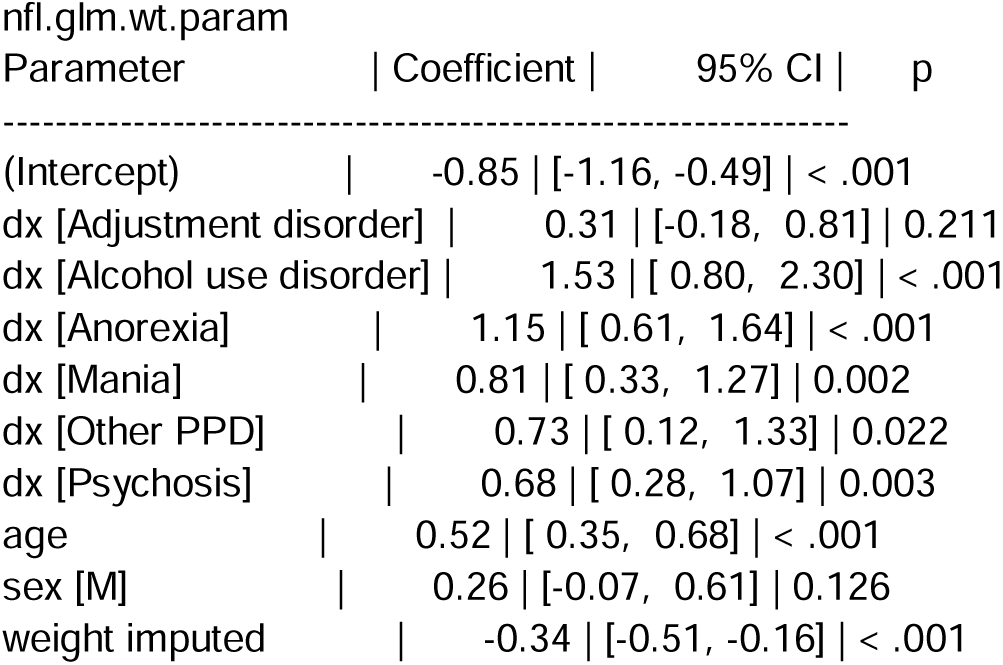
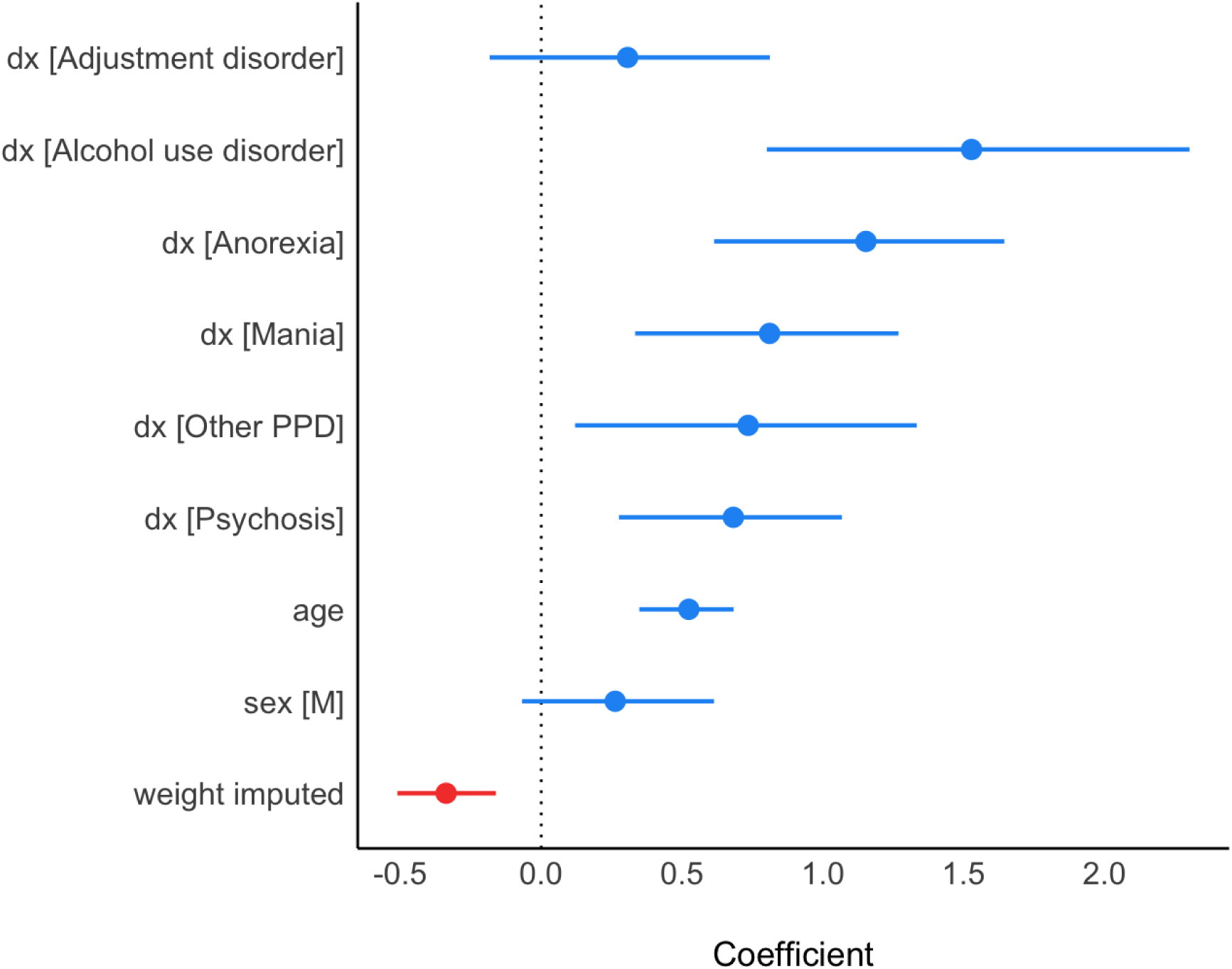

#### GFAP

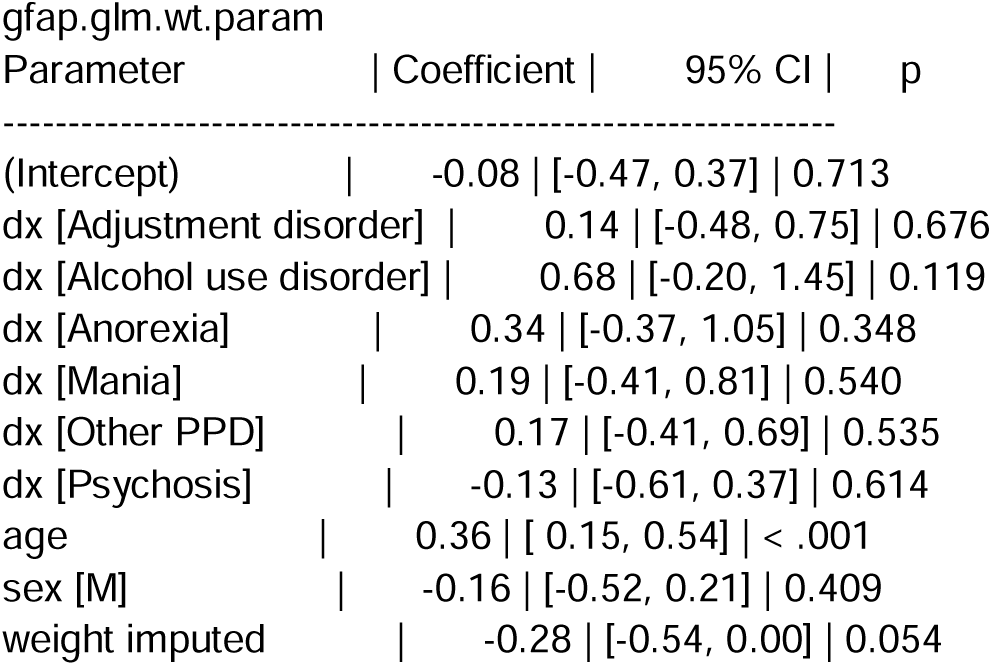
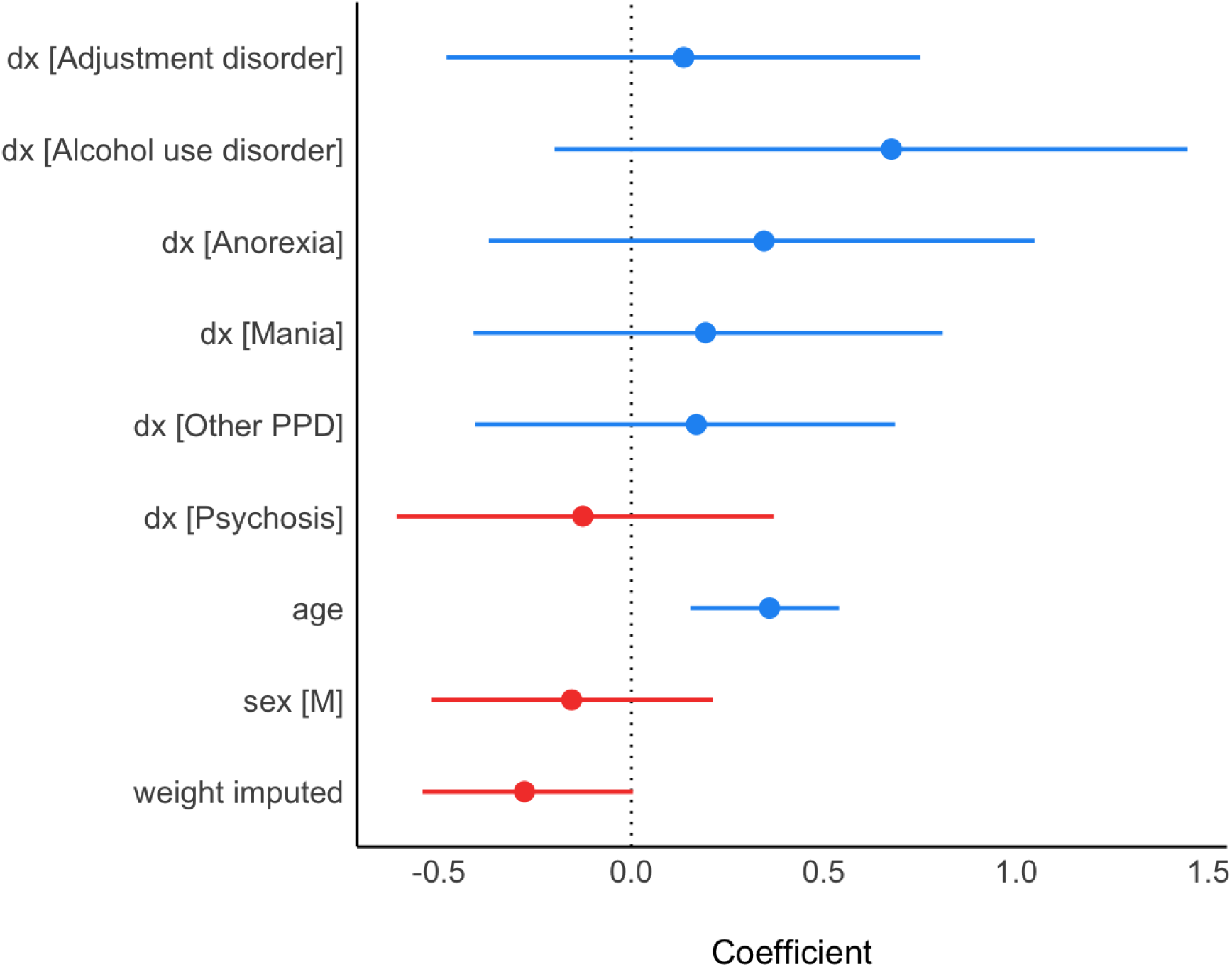

## References

Abé C, Ching CRK, Liberg B, et al. (2022) Longitudinal Structural Brain Changes in Bipolar Disorder: A Multicenter Neuroimaging Study of 1232 Individuals by the ENIGMA Bipolar Disorder Working Group. Biological Psychiatry 91(6): 582–592.

Abu-Rumeileh S, Abdelhak A, Foschi M, et al. (2023) The multifaceted role of neurofilament light chain protein in non-primary neurological diseases. Brain: A Journal of Neurology 146(2): 421–437.

Aggio V, Fabbella L, Finardi A, et al. (2022) Neurofilaments light: Possible biomarker of brain modifications in bipolar disorder. Journal of Affective Disorders 300: 243–248.

Al Shweiki MR, Steinacker P, Oeckl P, et al. (2019) Neurofilament light chain as a blood biomarker to differentiate psychiatric disorders from behavioural variant frontotemporal dementia. Journal of Psychiatric Research 113: 137–140.

Baudouin E, Duron E, Verdoux M, et al. (2025) Strong association between psychiatric disorders co-occurrence and dementia: a Bayesian approach on a 14-year clinical data warehouse. BMJ Mental Health 28(1): 1–7.

Bavato F, Cathomas F, Klaus F, et al. (2021) Altered neuroaxonal integrity in schizophrenia and major depressive disorder assessed with neurofilament light chain in serum. Journal of Psychiatric Research 140: 141–148.

Bavato F, Barro C, Schnider LK, et al. (2024) Introducing neurofilament light chain measure in psychiatry: current evidence, opportunities, and pitfalls. Molecular Psychiatry 29(8): 2543–2559.

Bavato F, Seifritz E and Quednow BB (2024) The multifaceted role of neurofilament light chain protein: emerging opportunities in primary psychiatric conditions. Brain: A Journal of Neurology 147(1): e5–e6.

Bousman CA, Luza S, Mancuso SG, et al. (2019) Elevated ubiquitinated proteins in brain and blood of individuals with schizophrenia. Scientific Reports 9(1): 2307.

Ceylan MF, Tural Hesapcioglu S, Kanoğlu Yüksekkaya S, et al. (2023) Changes in neurofilament light chain protein (NEFL) in children and adolescents with Schizophrenia and Bipolar Disorder: Early period neurodegeneration. Journal of Psychiatric Research 161: 342–347.

Cropley VL, Klauser P, Lenroot RK, et al. (2017) Accelerated Gray and White Matter Deterioration With Age in Schizophrenia. The American Journal of Psychiatry 174(3): 286–295.

Dai N, Jones BDM and Husain MI (2022) Astrocytes in the Neuropathology of Bipolar Disorder: Review of Current Evidence. Brain Sciences 12(11): 1513.

Doose A, Tam FI, Hellerhoff I, et al. (2023) Triangulating brain alterations in anorexia nervosa: a multimodal investigation of magnetic resonance spectroscopy, morphometry and blood-based biomarkers | Translational Psychiatry. Translational Psychiatry 13(1): 277.

Eratne D, Janelidze S, Malpas CB, et al. (2022) Plasma neurofilament light chain protein is not increased in treatment-resistant schizophrenia and first-degree relatives. The Australian and New Zealand Journal of Psychiatry 56(10): 1295–1305.

Eratne D, Kang MJY, Lewis C, Dang C, Malpas CB, Keem M, et al. (2024) Plasma and CSF neurofilament light chain distinguish neurodegenerative from primary psychiatric conditions in a clinical setting. Alzheimer’s & Dementia: The Journal of the Alzheimer’s Association 20(11): 7989–8001.

Eratne D, Kang M, Malpas C, et al. (2024) Plasma neurofilament light in behavioural variant frontotemporal dementia compared to mood and psychotic disorders. The Australian and New Zealand Journal of Psychiatry 58(1): 70–81.

Eratne D, Kang MJY, Lewis C, Dang C, Malpas C, Ooi S, et al. (2024) Plasma neurofilament light outperforms glial fibrillary acidic protein in differentiating behavioural variant frontotemporal dementia from primary psychiatric disorders. Journal of the Neurological Sciences 467: 123291.

Hellerhoff I, King JA, Tam FI, et al. (2021) Differential longitudinal changes of neuronal and glial damage markers in anorexia nervosa after partial weight restoration. Translational Psychiatry 11(1): 86.

Hou Y-C, Bavato F, Liu T-H, et al. (2025) Plasma neurofilament light chain levels are associated with delirium tremens in patients with alcohol use disorder. Progress in Neuro-Psychopharmacology & Biological Psychiatry 136: 111189.

Huang M-C, Tu H-Y, Chung R-H, et al. (2024) Changes of neurofilament light chain in patients with alcohol dependence following withdrawal and the genetic effect from ALDH2 Polymorphism. European Archives of Psychiatry and Clinical Neuroscience 274(2): 423–432.

Kang M, Grewal J, Eratne D, et al. (2025) Neurofilament light and glial fibrillary acidic protein in mood and anxiety disorders: A systematic review and meta-analysis. Brain, Behavior, and Immunity 123: 1091–1102.

Kang M, Eratne D, Dean O, et al. (2025) Plasma Glial Fibrillary Acidic Protein and Neurofilament Light Are Elevated in Bipolar Depression: Evidence for Neuroprogression and Astrogliosis. Bipolar Disorders. Epub ahead of print 23 April 2025. DOI: 10.1111/bdi.70029.

Kang MJY, Eratne D, Wannan C, et al. (2024) Plasma neurofilament light chain is not elevated in people with first-episode psychosis or those at ultra-high risk for psychosis. Schizophrenia Research 267: 269–272.

Light V, Jones SL, Rahme E, et al. (2024) Clinical Accuracy of Serum Neurofilament Light to Differentiate Frontotemporal Dementia from Primary Psychiatric Disorders is Age-Dependent. The American Journal of Geriatric Psychiatry: Official Journal of the American Association for Geriatric Psychiatry 32(8): 988–1001.

Livingston G, Huntley J, Liu KY, et al. (2024) Dementia prevention, intervention, and care: 2024 report of the Lancet standing Commission. Lancet (London, England) 404(10452): 572–628.

Lotan A, Luza S, Opazo CM, et al. (2023) Perturbed iron biology in the prefrontal cortex of people with schizophrenia. Molecular Psychiatry 28(5): 2058–2070.

Mostaid MS, Lee TT, Chana G, et al. (2017) Elevated peripheral expression of neuregulin-1 (NRG1) mRNA isoforms in clozapine-treated schizophrenia patients. Translational Psychiatry 7(12): 1280.

Nadler E, Kish SJ, Tong J, et al. (2025) Astrocyte alterations and dysfunction in alcohol use disorder: A comprehensive scoping review of clinical postmortem and preclinical evidence. Progress in Neuro-Psychopharmacology & Biological Psychiatry 142: 111509.

Nilsson IAK, Millischer V, Karrenbauer VD, et al. (2019) Plasma neurofilament light chain concentration is increased in anorexia nervosa. Translational Psychiatry 9(1): 180.

Queissner R, Buchman A, Demjaha R, et al. (2024) Serum neurofilament light as a potential marker of illness duration in bipolar disorder. Journal of Affective Disorders. Epub ahead of print 10 January 2024. DOI: 10.1016/j.jad.2024.01.088.

Requena-Ocaña N, Araos P, Serrano-Castro PJ, et al. (2023) Plasma Concentrations of Neurofilament Light Chain Protein and Brain-Derived Neurotrophic Factor as Consistent Biomarkers of Cognitive Impairment in Alcohol Use Disorder. International Journal of Molecular Sciences 24(2): 1183.

Rodrigues-Amorim D, Rivera-Baltanás T, Del Carmen Vallejo-Curto M, et al. (2020) Plasma β-III tubulin, neurofilament light chain and glial fibrillary acidic protein are associated with neurodegeneration and progression in schizophrenia. Scientific Reports 10(1): 14271.

Squitti R, Fiorenza A, Martinelli A, et al. (2024) Neurofilament Light Protein as a Biomarker in Severe Mental Disorders: A Systematic Review. International Journal of Molecular Sciences 26(1): 61.

Steinacker P, Al Shweiki MR, Oeckl P, et al. (2021) Glial fibrillary acidic protein as blood biomarker for differential diagnosis and severity of major depressive disorder. Journal of Psychiatric Research 144: 54–58.

Sun D, Stuart GW, Jenkinson M, et al. (2009) Brain surface contraction mapped in first-episode schizophrenia: a longitudinal magnetic resonance imaging study. Molecular Psychiatry 14(10): 976–986.

Wannan CMJ, Eratne D, Santillo AF, et al. (2024) Plasma neurofilament light protein is differentially associated with age in individuals with treatment-resistant schizophrenia and bipolar affective disorder compared to controls. Psychiatry Research 339: 116073.

Zarglayoun H, Jones SL, Light V, et al. (2025) Serum Neurofilament Light as a Neuropsychiatric Disorder Screening Test in Psychiatric Emergency Settings. The Journal of Neuropsychiatry and Clinical Neurosciences: appineuropsych20240228.

